# Outdoor attractive targeted sugar bait Phase III trials for malaria control in Kenya, Mali, and Zambia: An individual participant data meta-analysis

**DOI:** 10.64898/2026.05.06.26352614

**Authors:** Ruth A. Ashton, Daniel P. McDermott, Fousseyni Kane, Sophie Sarrassat, Angela Harris, Christen Fornadel, Joseph Wagman, Javan Chanda, Megan Littrell, Feiko O. ter Kuile, Aaron M. Samuels, Eric Ochomo, Thomas S. Churcher, Joseph Biggs, Sarah G. Staedke, Seydou Doumbia, Immo Kleinschmidt, Joshua Yukich, Thomas P. Eisele

## Abstract

Attractive targeted sugar baits (ATSB) are a potential new class of vector control tool that act through an “attract and kill” mechanism on mosquitoes. We conducted a meta-analysis using data from three large-scale Phase III trials of the Westham Sarabi v1.2 ATSB (0.11% dinotefuran) conducted in Kenya, Mali, and Zambia, to determine the effect of the intervention on clinical malaria incidence in children, *Plasmodium falciparum* infection prevalence, and dominant vector species parity, abundance, landing rate, and sporozoite positivity. The Sarabi ATSB was deployed on exterior walls at the rate of two per residential structure. Aggregated and individual-level meta-analyses were completed for each of the six trial outcomes, comprising 6981 person-years of follow-up for clinical malaria incidence (primary epidemiological outcome) and 19443 *Anopheles* for parity assessment (primary entomological outcome). Post-hoc analyses included assessment of dose-response relationships between coverage-adjusted intervention density and clinical malaria incidence. There were no statistically significant differences between arms in any of the epidemiological or entomological outcomes. There was statistically significant evidence of a 19% reduction in clinical malaria incidence for every 10 bait stations per hectare increase observed in spatial density (IRR 0.81, 95% CI 0.74-0.89, p<0.001), provided that the ATSB were in good condition. This finding suggests that there may be deployment approaches or dosing strategies under which ATSB tools could be efficacious, although threshold spatial densities could not be determined from available data. This meta-analysis furthermore highlights important recommendations for future cluster-randomized trials of vector control interventions, including conducting comprehensive baseline data collection to identify cluster outliers or sites with differences in vector bionomics, and collecting a limited set of entomological outcomes in all trial clusters to ensure an adequately powered and balanced analysis of entomological effects.

## Introduction

Stalling progress against malaria has increased the urgency to identify new and complementary tools to suppress malaria transmission and protect individuals living in the highest burden areas. Attractive Targeted Sugar Bait (ATSB) stations are among several new vector control tools that have recently undergone large-scale field testing to explore their potential public health benefits. ATSBs take advantage of mosquitoes’ natural sugar feeding behavior, by providing an alternative source of sugar laced with an ingestion toxicant that induces mosquito mortality mosquito.

Early field studies of the ATSB paradigm tested the effect of spraying vegetation with a sugar-toxicant solution, or use of improvised sugar-toxicant delivery systems inside residential structures on mosquito density and landing rates [1–4]. Proof-of-principle studies were largely conducted in arid or semi-arid settings in Israel and Mali, where target mosquitoes were *An. sergentii, An. clavigeri*, or *An. gambiae s.l.* [1, 2, 5]. The Sarabi ATSB station (version 1.2, Westham Co., Hod-Hasharon, Israel) comprises date paste as an attractant and feeding stimulant and dinotefuran (0.11% w/w) as the active ingredient, behind a protective membrane which allowed feeding by mosquitoes but prevents most non-target organisms accessing the bait. A 14-village randomized trial in Mali where an earlier version of the Sarabi ATSB station was installed on exterior walls of residential structures indicated that the ATSB was associated with large reductions in abundance of *An. gambiae s.l.* and sporozoite prevalence [6]. These promising findings were also supported by modelling studies indicating that high mosquito ATSB feeding rates in areas of ITN use would lead to noticeable reductions in transmission [7–9]. Feeding studies deploying prototype Sarabi bait stations without toxicant on exterior walls indicated daily vector feeding rates over 25% for *An. gambiae s.l.* in Mali, and of 9% and 4.8% for *Anopheles funestus s.l.* in Zambia and Kenya, respectively, and no difference in feeding rates when two or three ATSB were deployed per structure [10–12]. The ATSB paradigm was initially proposed to the World Health Organization in 2014 [13]. To assess if outdoor use of the Westham Sarabi ATSB station demonstrated a public health benefit when deployed alongside existing vector control, large cluster-randomized controlled trials were completed in three different settings from 2021-2024 [14], with the aim to move rapidly to establishment of ATSB as a new vector control product class by the World Health Organization.

Generating the evidence required for WHO recommendation necessitated rigorous cluster-randomized trials (cRCTs), since the hypothesized mechanism of action involves area-wide vector population suppression that would confer community-level protection, precluding individual-level randomization. In cluster-randomized trials the unit of randomization is a group (i.e. village) rather than an individual, and outcomes are measured among members of the groups (i.e. village residents). Since measures of outcomes among group members cannot be assumed to be independent, as is the case in an individually-randomized trial, special methods must be applied in the design and analysis of cluster-randomized trials [15]. Over time, cRCTs have undergone methodological refinements such as the use of restricted randomization to improve baseline balance of covariates between arms [16, 17] and use of buffer areas around clusters to reduce between cluster contamination [15]. Furthermore, cRCTs allow quantification of both direct (i.e. for the user) and indirect (i.e. community-level) effects of interventions such as vector control tools.

In this paper we document findings from pooled and individual-level meta-analysis of the three Phase III trials of outdoor use of the Westham Sarabi ATSB against epidemiological and entomological outcomes, together with a secondary analysis exploring if ATSBs are more effective when deployed at higher spatial densities. Secondly, we present key lessons learned from the design, implementation, and analysis of these trials along with recommendations for future cRCT of vector control interventions.

## Materials and methods

### Study selection criteria and summary of eligible studies

This meta-analysis was limited to the three Phase III trials of the Sarabi version 1.2 ATSB to evaluate efficacy against epidemiological and entomological outcomes. Due to differences in active ingredient, delivery mechanism (e.g. foliage spray, improvised or manufactured bait station) and deployment location (e.g. indoors or outdoors), studies evaluating ATSB devices other than Sarabi version 1.2 were excluded. Additionally, no other studies have evaluated epidemiological outcomes of ATSB deployment.

The three Phase III ATSB trials followed a master protocol and statistical analysis plan [14, 18], but were conducted independently in Mali, Kenya, and Zambia from 2021 to 2024. The trials were approved by local ethics committees at each site and international partner institutional review boards, and primary findings have been published for each site [19–22]. While intent to conduct meta-analysis was stated in the published trial statistical analysis plan [18], the meta-analysis was not registered and a formal meta-analysis protocol was not developed. PRISMA-IPD guidance were followed in reporting findings [23]. The objective was to conduct pooled and individual participant data meta-analyses for the primary and secondary outcomes, then to conduct further exploratory analyses to guide future research hypotheses.

Individual-level anonymized data were standardized across sites and combined into a single file for each outcome. Since the three trials followed a master protocol, all individuals included for the site-specific primary analyses were retained for meta-analyses of individual participant data (IPD). For epidemiological outcomes, this entailed a child-level summary for each cohort participant and individual-level survey participant outcomes. For entomological outcomes, trap-level data were used for abundance and landing rate outcomes, and mosquito-level data for parity and sporozoite testing results. Additionally, data describing ATSB density and coverage were collated across the three sites.

In the three trials, Sarabi version 1.2 ATSB bait stations (hereafter referred to as ATSB) were deployed on exterior walls of domestic structures, applying two bait stations per structure. The intervention was deployed for two transmission seasons in Zambia (8 months each season). In Mali and Kenya ATSB were deployed continuously, with all devices replaced every eight or six months, respectively, and total deployment period of 21 months in Mali and 24 months in Kenya. In addition to replacement campaigns in which all ATSB were replaced with new devices, the intervention was subject to periodic monitoring during which those found to be missing or damaged were replaced. ATSB were deployed alongside the national standard of care vector control (universal ITN coverage in Kenya and Mali, either ITN or IRS in Zambia).

### Data analysis

The primary outcome of the three trials was clinical malaria incidence among cohorts of children in each study cluster (age 1-14 years in Kenya and Zambia, age 5-14 years in Mali). The clinical case definition differed by site in the confirmatory test required alongside reported or measured fever: either positive RDT or microscopy slide in Kenya, both positive RDT and microscopy slide in Mali, and positive RDT in Zambia (Table 1). Secondary outcomes included *P. falciparum* infection prevalence among individuals aged six months or older (estimated by HRP2 RDT in Kenya and Zambia, and by HRP2-pLDH RDT in Mali), and entomological indicators assessed among the primary vectors at each site. The entomological indicators were: parity, abundance, landing rate, sporozoite positivity [14]. The trials were powered to detect a ≥30% reduction in clinical malaria incidence among children, a ≥30% reduction in *P. falciparum* prevalence, and an increase in the dominant vector species proportion non-parous (i.e. % of mosquitoes that have never laid eggs) of 9 percentage points (from 48.8% to 57.8%), equivalent to a relative increase of 18% in the proportion non-parous [18]. Entomological outcomes were assessed in a subset of trial clusters: in Kenya a stratified simple random sample was used to select 16 clusters, 8 per subcounty; in Mali 28 clusters were selected by simple random sample; in Zambia a purposive approach was used to select 20 clusters with a similar number of clusters per district but excluding areas with accessibility challenges, known low A*nopheles* abundance, or community acceptance concerns relating to entomological surveillance.

**Table 1:**
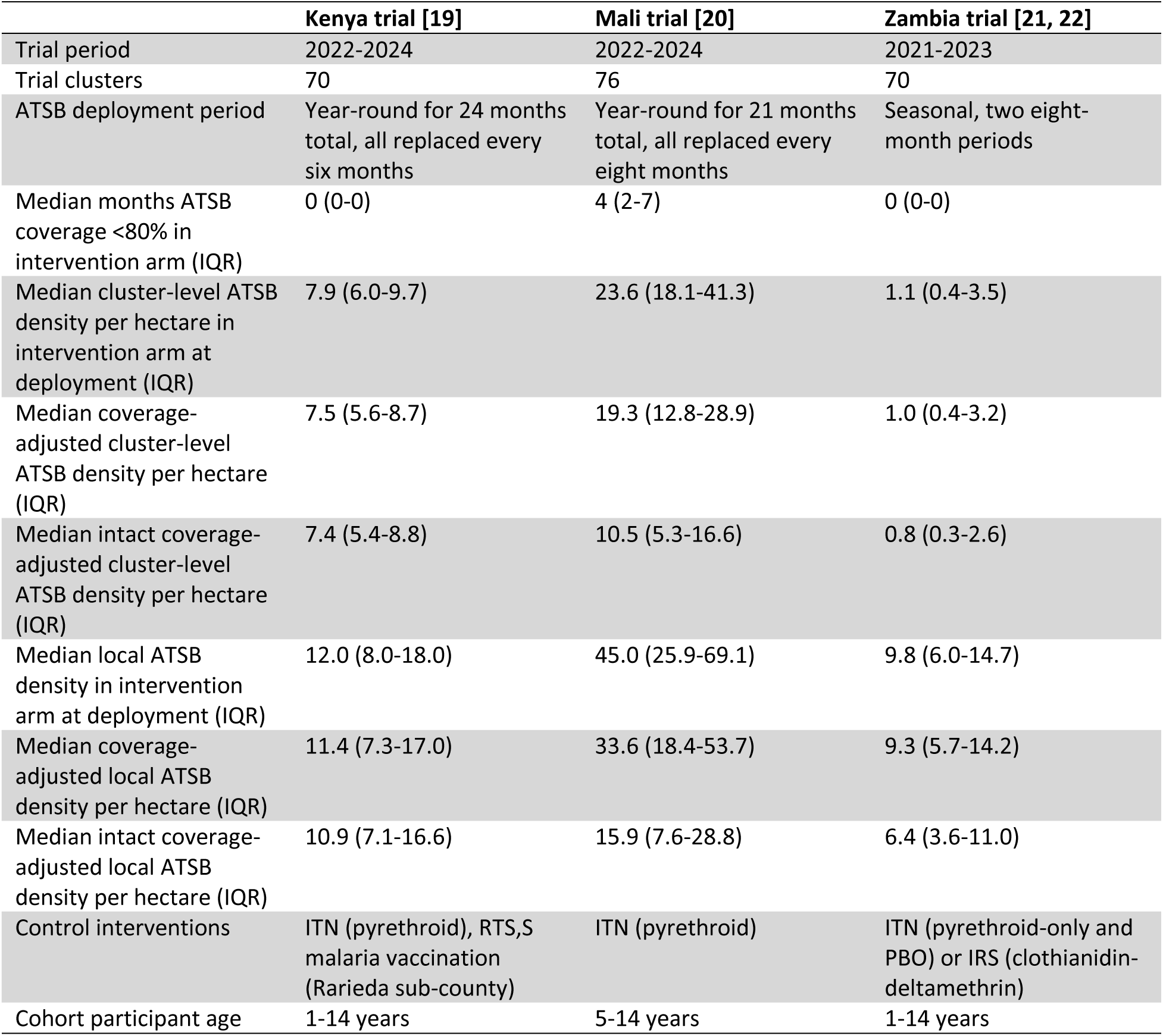
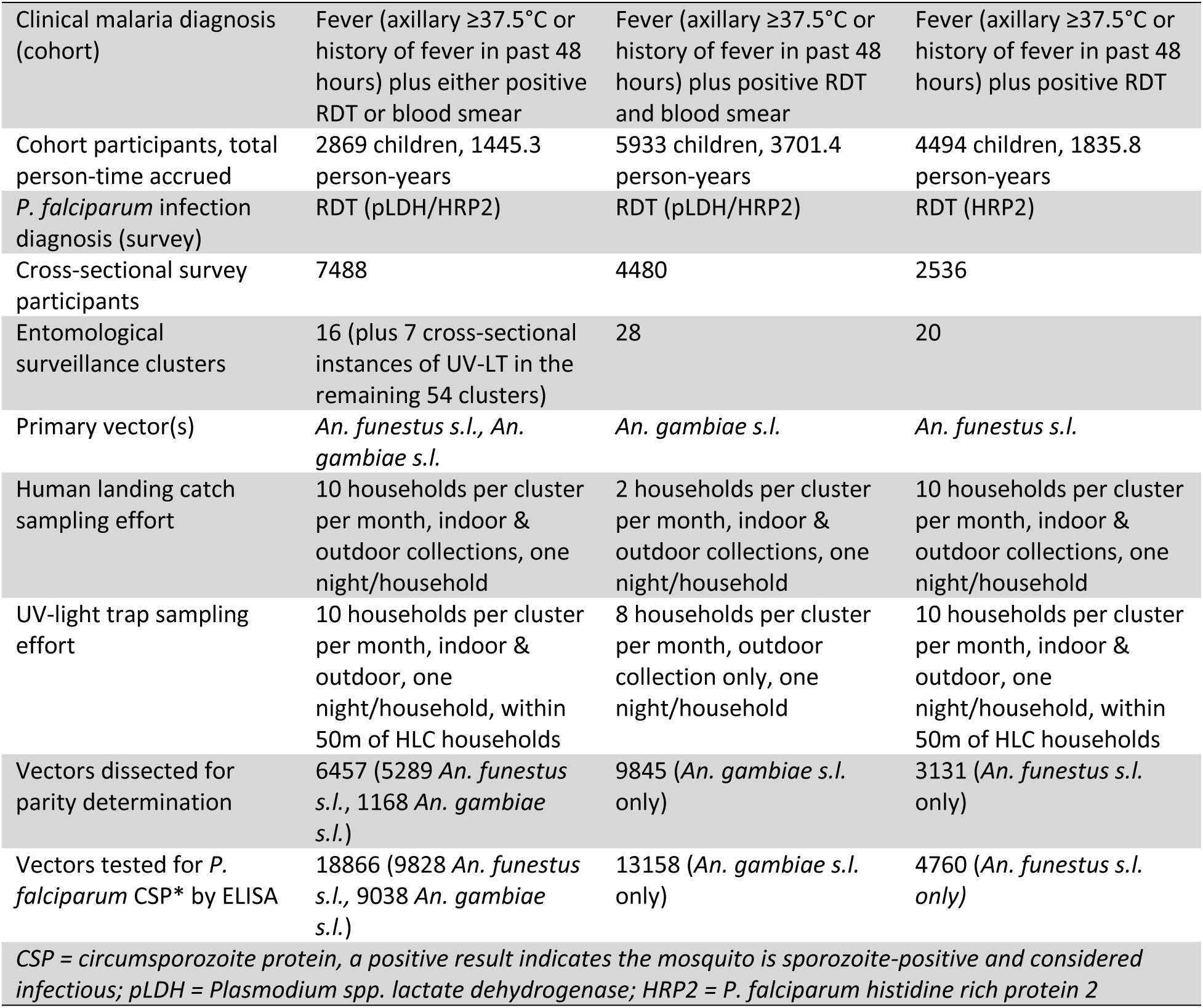
Characteristics of the Phase III Westham Sarabi v1.2 ATSB trials completed in Kenya, Mali, and Zambia.

A pooled meta-analysis used site-level efficacy results for primary and secondary outcomes as reported in previous publications [19–22]. Entomological outcomes from the Mali site were not published at the time of this analysis, consequently a two-stage approach was used whereby efficacy estimates for each entomological outcome were estimated for the Mali site, then included in the pooled meta-analysis. IPD analyses were conducted using the intention-to-treat population, where models included country as a fixed effect and trial cluster (unique across sites) as a random effect. Models used a Poisson structure for count outcomes (clinical incidence, vector abundance, landing rate), and a Binomial structure for binary outcomes (parity, sporozoite positivity). Sensitivity analyses using negative binomial model structure gave similar results to those from Poisson structure models.

Covariables tested in adjusted IPD models included age, sex, reported ITN use over all follow-up visits (never, sometimes, always), household socioeconomic status, household construction, number of household members, local ATSB density (number ATSB deployed within 1 hectare area estimated as a circle with 56.4m radius around participant sleeping structure), cluster-level ATSB density (number ATSB deployed in cluster divided by cluster area in hectares), cluster-level density of residential structures, ATSB coverage (proportion of structures with at least two ATSB installed on exterior walls), and coverage of ATSB in good condition (proportion of structures with at least two ATSB that do not meet pre-defined damage criteria [24] installed on exterior walls). ATSB coverage was assessed in Mali and Zambia during monthly cohort follow-up visits, and in Kenya during bimonthly visits to all intervention cluster households. Cluster means of monthly total rainfall from CHIRPS 3.0 at 0.05 degree resolution [25], and monthly enhanced vegetation index (EVI) from MODIS at 1km resolution [26] were generated. Land use classification in 2020 from Landsat at ∼30 meter resolution [27] was extracted for each participant location. The master protocol defined target ATSB coverage as 100%, but did not set a minimum threshold for acceptable coverage [14, 18]. For post-hoc per protocol IPD analysis, we excluded intervention arm clusters where ATSB coverage was <80% for 6 months or more.

To further assess any relationships between ATSB density and clinical malaria incidence, dose-response analyses were conducted. Six exposure indicators were tested in models: cluster-level ATSB density; coverage-adjusted cluster ATSB density (the product of cluster-level ATSB density and mean coverage of ATSB in any condition in the cluster); intact coverage-adjusted cluster ATSB density (the product of cluster-level ATSB density and mean coverage of ATSB in the cluster that did not require replacement); local ATSB density; coverage-adjusted local ATSB density; intact coverage-adjusted local ATSB density. Models included one of these six exposure indicators in place of trial arm, along with baseline cluster incidence and country as fixed effects, and cluster as a random effect. Models were developed both including and excluding control arm clusters, and using either the full range of densities observed or the range of shared support (range of individual observations that existed in all three sites, supplementary table 1).

Further post-hoc analyses included classifying clusters by the most common vector morphologically identified, exploring evidence for ATSB effect modification by vector species. In Mali and Zambia this analysis was limited to the 28 and 20 clusters, respectively, where entomological surveillance was conducted. In Kenya, seven rounds of UV light trap collections were completed (one at baseline and six during trial implementation, each round consisting of ten households per cluster), enabling vector classification across all 70 trial clusters.

To assess between-cluster heterogeneity, the incidence coefficient of variation (CV) was calculated for each site’s baseline data (clinical incidence cohort in all three sites and cross-sectional survey in Mali and Zambia), and by arm during the trial using two methods. First, following the method of moments approach specified by Hayes and Moulton, whereby cluster summaries are used to generate a point estimate of CV for each dataset [15]. Second, using a linear mixed effects model without predictors to generate a point estimate and 95% confidence interval for CV [28]. The CV by arm during the trial was also generated for all secondary outcomes: prevalence, primary vector parity, abundance, landing rate, and sporozoite positivity (supplementary table 2).

All meta-analyses were completed in R version 4.2.2. Pooled meta-analyses used the metafor package and IPD analyses used the lme4 package. Model-based estimation of coefficient of variation was performed in Stata version 14. Due to inclusion of only three studies in the meta-analysis, formal assessment of publication bias was not conducted.

### Ethics statement

The trials included in this meta-analysis were approved by local ethics committees at each site and international partner institutional review boards. The Kenya trial was approved by the Kenya Medical Research Institute Scientific and Ethics Review Unit (4189), the Liverpool School of Tropical Medicine Ethics Committee (21-027), and US Centers for Disease Control and Prevention (00008118) via a reciprocal agreement with KEMRI SERU. The Mali trial was approved by Institutional Review Boards at the University of Bamako (2021/124/CE/USTTB) and the London School of Hygiene and Tropical Medicine (17283-1). The Zambia trial was approved by the University of Zambia Biomedical Research Ethics Committee (1197-2020), PATH Research Ethics Committee (1460046-5) and Tulane University Biomedical Institutional Review Board (2019-595). All three trials were registered on Clinicaltrials.gov: Kenya NCT05219565, Mali NCT04149119, Zambia NCT 04800055.

## Results

The Phase III trials of Westham Sarabi v1.2 ATSB in Kenya, Mali and Zambia included a total of 6981 person-years of follow-up time accrued among 13,296 children participating in the cohort study, with 9157 incident malaria episodes recorded. The trials also included a total of 14504 household survey participants (Table 1). All requested IPD data from the three trial sites were obtained for analysis, no data quality or completeness issues were identified.

Pooled meta-analysis indicates that ATSB deployment was associated with an overall 9% reduction in clinical malaria incidence among children (IRR 0.91, 95% CI 0.82-1.02) and an overall 7% reduction in *P. falciparum* infection prevalence in community members aged six months and older (OR 0.93, 95% CI 0.80-1.10) (Figure 1).

**Figure 1:** Effect of Sarabi v1.2 ATSB on clinical malaria incidence among children (top) and *P.* falciparum infection prevalence among community members aged ≥6 months (bottom), assessed by pooled meta-analysis using crude site-specific effect estimates.

Pooled meta-analyses of entomological outcomes indicate that ATSB deployment was associated with increased parity (i.e. older age) of female mosquitoes (OR 1.36, 95% CI 0.89-2.06) and significantly increased odds of being sporozoite positive (OR 1.23, 95% CI 1.01-1.49), while both abundance of primary vectors assessed by UV light trap and nightly landing rate of primary vectors were slightly reduced in clusters where ATSB were deployed (IRR 0.73, 95% CI 0.44-1.24, and IRR 0.73, 95% CI 0.41-1.30, respectively) (Figure 2). Effect estimates of ATSB on both epidemiological and entomological outcomes from the pooled meta-analysis had low certainty as indicated by wide 95% confidence intervals.

**Figure 2:** Effect of Westham Sarabi v1.2 ATSB on primary vector parity (top), abundance (second plot), landing rate (third plot), and sporozoite positivity (bottom), assessed by pooled meta-analysis using crude site-specific effect estimates. Data are reported among *An. gambiae s.l.* and *An. funestus s.l.* in Kenya, among *An. gambiae s.l.* only in Mali, and among *An. funestus s.l.* only in Zambia.

Findings from individual participant data analyses of epidemiological outcomes indicated smaller reductions in incidence (IRR 0.97, 95% CI 0.84-1.12) and prevalence (OR 0.93, 95%CI 0.76-1.14) than the pooled model results, and with wider 95% confidence intervals (Table 2). Restricting analysis to a per-protocol population, in which intervention arm clusters that experienced more than six months of ATSB coverage (proportion of visited structures with fewer than two ATSB installed) below 80% were excluded did not substantially change the findings (Table 2).

**Table 2:** Summary of individual participant data models from Kenya, Mali and Zambia sites combined, describing effect of ATSB on the defined outcomes. Intention-to-treat models included data from all clusters, while per-protocol models excluded any intervention arm clusters where ATSB coverage (proportion of residential structures with <2 ATSB) was below 80% for more than 6 months. The ITT analyses for epidemiological outcomes included 108 clusters per arm, while PP analyses excluded 15 ATSB arm clusters. The ITT analyses for entomological outcomes included data from 32 clusters per arm, while PP analyses excluded 7 ATSB arm clusters.

| Outcome | Populatio<br>n | Crude model |  |  | Adjusted model* |  |  |
| --- | --- | --- | --- | --- | --- | --- | --- |
|  |  | N<br>cluster<br>s | Effect estimate<br>(95% CI) | p | N<br>clusters | Effect estimate<br>(95% CI) | p |
| Clinical incidence<br>(primary) | ITT | 216 | 0.97 (0.83-1.13) | 0.663 | 213 | 0.97 (0.84-1.12) | 0.702 |
|  | PP | 201 | 0.95 (0.81-1.12) | 0.576 | 198 | 0.96 (0.82-1.12) | 0.557 |
| Prevalence | ITT | 216 | 0.94 (0.76-1.17) | 0.573 | 213 | 0.93 (0.76-1.14) | 0.493 |
|  | PP | 201 | 0.96 (0.76-1.21) | 0.730 | 198 | 0.92 (0.74-1.14) | 0.443 |
| Parity<br>(primary) | ITT | 64 | 1.54 (0.91-2.60) | 0.109 | 63 | 1.44 (0.83-2.48) | 0.191 |
|  | PP | 57 | 1.34 (0.80-2.24) | 0.265 | 56 | 1.25 (0.73-2.12) | 0.417 |
| Abundance | ITT | 64 | 0.80 (0.49-1.29) | 0.350 | 63 | 0.82 (0.50-1.36) | 0.444 |
|  | PP | 57 | 0.80 (0.47-1.39) | 0.441 | 56 | 0.83 (0.47-1.47) | 0.525 |
| Landings | ITT | 64 | 0.77 (0.43-1.39) | 0.387 | 63 | 0.80 (0.44-1.44) | 0.455 |
|  | PP | 57 | 0.74 (0.37-1.49) | 0.395 | 56 | 0.82 (0.43-1.55) | 0.534 |
| Sporozoite positive | ITT | 64 | 1.17 (0.91-1.51) | 0.223 | 63 | 1.13 (0.87-1.46) | 0.368 |
|  | PP | 57 | 1.19 (0.90-1.56) | 0.228 | 56 | 1.14 (0.86-1.51) | 0.380 |
| ITT=Intention-to-treat. PP=Per-protocol. Pop=population.<br>*Baseline incidence observation was unavailable for 3 clusters in Zambia. Adjusted model covariates are baseline incidence, age, sex, improved roof (binary) and improved (wall) for incidence outcome; baseline incidence, age and sex for prevalence outcome; baseline incidence, indoor/outdoor trap location and rainfall in previous month for parity outcome; baseline incidence and rainfall in previous month for abundance outcome; baseline incidence, indoor/outdoor location and % of cluster cohort participants reporting always sleeping under an ITN for landings outcome; and baseline incidence, collection method (UV light trap or human landing collection) and rainfall in previous month for sporozoite positivity outcome |  |  |  |  |  |  |  |

Results from individual-level models of entomological outcomes were found to be similar to the pooled model results, suggesting a slight but not statistically significant increase in proportion parous and sporozoite positivity in the intervention arm. Restricting the parity outcome model to the per-protocol population resulted in a smaller effect size than in the ITT population, but still suggestive of an increase in mosquito age in the intervention arm. Trap-level models with abundance and landing rate outcome indicated similar results to the pooled model, suggesting small declines in abundance and landing rate in the ATSB arm but with wide confidence intervals, consistent in both crude and adjusted models and intention-to-treat and per-protocol populations.

ATSB density varied widely between sites (Table 1) due to differences in settlement patterns, with the lowest density in Zambia and the highest in Mali. In Kenya, coverage- and intact coverage-adjusted ATSB densities were similar to observed densities (Table 1). In Mali, coverage- and intact coverage-adjusted ATSB densities were lower than observed densities due to both reduced coverage and high damage rates among those ATSB present. In Zambia, intact coverage-adjusted densities were slightly reduced due to high damage rates in some clusters.

There was consistent evidence for reductions in malaria incidence associated with increasing cluster-level ATSB density after accounting for coverage (multiplying density by mean % cluster coverage). Increasing cluster-level ATSB density by 10 (after accounting for coverage of ATSB in good condition) resulted in 19% reduction in clinical malaria incidence (IRR 0.81, 95% CI 0.74-0.89, p<0.001). A smaller effect size was observed when adjusting cluster-level ATSB density by coverage of ATSB in any condition (IRR 0.93, 95% CI 0.88-1.00, p=0.041), suggesting that efficacy of ATSBs is likely to be contingent on their physical condition. Models examining coverage-adjusted local ATSB density as the exposure similarly found evidence for an association with reduced clinical malaria incidence (Table 3), but with a smaller effect size per additional 10 ATSBs per hectare. There was no evidence for a dose-response relationship between continuous cluster or local ATSB density indicators and clinical malaria incidence when density measures were not adjusted for ATSB coverage (Table 3).

**Table 3:** Effect of ATSB density on malaria clinical incidence among children. Models include baseline cluster incidence and site as fixed effects, and cluster as random effect. Cluster-level density was defined as the total ATSB deployed in the cluster divided by the total area of the cluster in hectares. Local density was defined as the number of ATSBs deployed in a 1-hectare area surrounding the cohort participant household. In control arm clusters, ATSB density indicator values were 0

| Density | Population | Exposure* | N children / clusters | IRR | 95% CI | p |
| --- | --- | --- | --- | --- | --- | --- |
| Cluster-level | ATSB and control arms | Cluster-level density/10 | 13118 / 213 | 0.96 | 0.91-1.02 | 0.154 |
|  |  | Coverage-adjusted cluster-level density/10 | 13118 / 213 | 0.93 | 0.88-1.00 | 0.041 |
|  |  | Intact coverage-adjusted cluster-level density/10 | 13118 / 213 | 0.81 | 0.74-0.89 | <0.001 |
|  | ATSB arm only | Cluster-level density/10 | 6565 / 106 | 0.94 | 0.86-1.02 | 0.135 |
|  |  | Coverage-adjusted cluster-level density/10 | 6565 / 106 | 0.91 | 0.83-0.99 | 0.025 |
|  |  | Intact coverage-adjusted cluster-level density/10 | 6565 / 106 | 0.76 | 0.68-0.85 | <0.001 |
| Local | ATSB and control arms | Local density/10 | 12601 / 213 | 0.99 | 0.97-1.01 | 0.407 |
|  |  | Coverage-adjusted local density/10 | 12601 / 213 | 0.99 | 0.97-1.02 | 0.576 |
|  |  | Intact coverage-adjusted local density/10 | 12601 / 213 | 0.96 | 0.93-1.00 | 0.029 |
|  | ATSB arm only | Local density/10 | 6048 / 106 | 0.99 | 0.97-1.01 | 0.536 |
|  |  | Coverage-adjusted local density/10 | 6048 / 106 | 1.00 | 0.97-1.02 | 0.715 |
|  |  | Intact coverage-adjusted local density/10 | 6048 / 1.06 | 0.96 | 0.93-1.00 | 0.029 |

*\*Models are fitted using rescaled density values which have been divided by 10, effect estimates therefore reflect effect of a 10-unit increase in ATSB density on malaria incidence*

Post-hoc analysis exploring the effect of ATSB deployment on clinical malaria incidence in the subset of clusters which completed monthly entomological surveillance suggested an enhanced effect in crude models (IRR 0.76, 95% CI 0.58-0.98), but inclusion of baseline cluster incidence in the model resulted in widened confidence intervals for this effect (IRR 0.84, 95% CI 0.39-1.79). Inspection of data suggests that among the clusters conducting entomology surveillance collections, baseline malaria incidence was imbalanced across the arms, with average higher incidence in the intervention arm.

In post-hoc analysis exploring evidence for effect modification by primary vector, all 28 entomological surveillance clusters in Mali and 52 of the total 70 clusters in Kenya had *An. gambiae s.l.* as the primary vector. In all 20 entomological surveillance clusters in Zambia, and the remaining 18 clusters in Kenya, *An. funestus s.l.* was the primary vector. Findings from individual-level meta-analysis did not vary substantially when restricting models to outcomes among either *An. gambiae s.l.* or among *An. Funestus s.l.* (data not shown). When assessing the data from Kenya only, where all 70 trial clusters could be classified by primary vector species, there was evidence for effect modification. Subgroup analysis indicated that among clusters where *An. funestus* was predominant, clinical malaria incidence was higher in the intervention than control arm (IRR 1.71, 95% CI 1.08-2.70, p=0.021, based on 13 intervention clusters and 5 control clusters), while the ATSB effect estimate in the clusters where *An. gambiae* was predominant was IRR 0.76 (95% CI 0.48-1.20, based on 22 intervention clusters and 30 control clusters). The *An. funestus* predominant clusters in Kenya were largely located around Lake Kanyaboli, an area that includes a large papyrus swamp, and was characterized by very high clinical malaria incidence during the trial.

Estimates of ATSB efficacy in this meta-analysis all reported wide 95% CI, indicating substantial heterogeneity. Table 4 presents the estimates of coefficient of variation estimated for clinical incidence outcome at each of the three trial sites from different stages of the trial. Estimates of CV for prevalence and all entomological outcomes are shown in Supplementary Table 2. While the coefficient of variation was relatively consistent over time in Mali, the Kenya site reported a very high coefficient of variation during the trial, indicating very high heterogeneity between clusters. The Zambia site had very high CV at baseline due to limited follow-up time accrued, with estimated CV during the trial similar to the protocol estimate.

**Table 4:** Summary of coefficient of variation calculated by either Method-of-moments [15], or using a Poisson model with cluster-level random effect and no predictors pooling data from both trial years within each site [28].

| Estimate | Method-of-moments |  |  | Model-based estimate (95% CI) |  |  |
| --- | --- | --- | --- | --- | --- | --- |
|  | Kenya | Mali | Zambia | Kenya | Mali | Zambia |
| Protocol | 0.40 | 0.40 | 0.40 | N/A | N/A | N/A |
| Baseline* | 0.60 | 0.39 | 1.00 | 0.56<br>(0.39-0.81) | 0.38<br>(0.30-0.48) | 1.02<br>(0.34-1.40) |
| Trial, control arm | 0.75 | 0.36 | 0.35 | 0.75<br>(0.59-0.95) | 0.32<br>(0.24-0.42) | 0.40<br>(0.31-0.51) |
| Trial, intervention arm | 0.77 | 0.28 | 0.46 | 0.75<br>(0.59-0.95) | 0.27<br>(0.20-0.37) | 0.48<br>(0.38-0.61) |

\*Baseline CV estimates excluded any clusters that were dropped between baseline and trial. Three clusters in Zambia did not have any valid incidence data and are excluded from baseline CV estimation

## Discussion

This meta-analysis did not find any definitive evidence that deployment of two Westham Sarabi v1.2 ATSB on exterior walls of residential structures was associated with public health relevant reductions in clinical malaria incidence in children, *P. falciparum* infection prevalence, or entomological indicators including primary vector parity, sporozoite positivity, human landing rate, or abundance estimated from UV light traps. Dose-response analyses demonstrated that higher deployment densities of ATSB significantly reduce clinical malaria incidence (after accounting for variations in coverage and condition of ATSB), aligning with results from the Mali trial, which found a 26% reduction in clinical incidence comparing clusters with >80% coverage of ATSB in good condition to control clusters [20]. These results suggest further studies to define the required dosage of ATSB for an effect or assess alternative deployment strategies may be merited if an improved ATSB product with improved attractiveness, mosquito feeding rates, and physical durability becomes available.

Early proof-of-concept studies assessing the ATSB paradigm were conducted in Mali and Israel, arid environments with differences in vegetation and likely natural sugar availability compared to the trial sites in Kenya and Zambia. While dose-ranging or dose-finding studies with epidemiological outcomes were not conducted prior to the Phase III trials, early studies indicated that deployment of a prior version of the Sarabi ATSB was associated with reduced vector abundance and sporozoite positivity [6, 29]. Studies assessing vector feeding on modified Sarabi devices that did not include an active ingredient were targeted to locations within the Zambia trial sites that had relatively higher residential structure density than that observed in the Phase III trials [11], although the equivalent study in Kenya was conducted at sites broadly representative of the cRCT trial area [12]. The three trials were conducted in rural settings: if a high density of ATSB devices is required for effectiveness, it is possible that this intervention is more suited to peri-urban or urban areas with high structure and population density. The median cluster-level ATSB densities at the Kenya and Zambia sites were 8 and 1 per hectare, respectively, consequently increasing ATSB density by 10 per hectare is likely not feasible in these settings unless alternative deployment approaches are used. A recent study evaluating the effect of Westham Sarabi v1.2 ATSB on entomological outcomes in a Nigerian camp for internally displaced people was able to achieve high spatial density due to the highly clustered layout of the camp and deployment of three devices per house (one indoor and two outdoor) [30].

There remain multiple evidence gaps regarding this new paradigm, including but not limited to further understanding of *Anopheles* sugar-finding and sugar-feeding behavior, optimal deployment strategies and required spatial density of devices, and the characteristics of an optimal device for delivery of oral active ingredients in an “attract and kill” paradigm. Despite efforts to develop the Phase III trials of Westham Sarabi ATSB with independent expert review, and to rigorously implement the protocol, our findings highlight challenges and lessons learned that can inform the design of future vector control cRCTs (Table 5).

**Table 5:** Key recommendations for future cRCT of vector control interventions.

| Recommendation | Reasoning |
| --- | --- |
| Where there are limited prior efficacy data, consider conducting trials sequentially rather than in parallel | • Parallel trials may establish the evidence base to allow rapid scaling of new intervention, provided they are found efficacious<br>• Sequential trials permit findings from early trials to guide protocol amendments in subsequent trials, reducing risks of multiple trials with null result, however, this lengthens the timeline to eventual WHO recommendation and eventual scale-up. |
| Consider if site characteristics of proof-of-concept and/or Phase II studies will be matched across sites selected for Phase III sites | • Initial small-scale studies were designed to measure entomological outcomes and selected clusters to facilitate entomological surveillance, while Phase III trial clusters were designed to optimize epidemiological outcome measurement.<br>• Design criteria for Phase III trial clusters should consider if any characteristics of previous study sites are critical to replicate, such as ecological context (e.g. natural sugar availability) and housing density |
| Use hypothesis-driven approaches to selecting entomology outcomes | • By focusing on outcomes required to confirm intervention effect, mechanism of action or contextualize epidemiological outcomes, entomology surveillance costs can be minimized compared to the inclusion of multiple trapping methods and an exploratory approach to analysis |
| When feasible, assess key entomology outcomes in all trial clusters | • Limiting some outcomes to a subset of clusters results creates challenges in achieving balance between arms in the subset of clusters with entomological outcomes<br>• Subgroup analyses are at risk of Type 1 error |
| Ensure sufficient baseline data are collected to accurately estimate outcomes at baseline in all clusters | • Generates site-specific parameters to confirm sample size assumptions<br>• Where incidence data are collected, baseline data collection must accrue sufficient person-time (at least one full transmission season) to accurately estimate incidence, avoiding skew towards zero-incidence that results from limited follow-up time<br>• If an entomological outcome will be included in the trial, vector bionomics should be assessed in all clusters to confirm balance between arms and allow inclusion of entomological variables in restricted randomization |
| Conduct pilot and/or baseline data collection in sufficient clusters to permit exclusion of outlier clusters | • By conducting baseline data in excess clusters, any clusters with unexpected or outlier results can be excluded.<br>• The opportunity to exclude outliers may assist in controlling coefficient of variation and maximizing power, assuming that baseline outcomes are predictive of cluster-level outcomes during the trial |
| Include a “worst case scenario” cluster-level power calculation in the protocol, to determine the ability to assess outcomes if cluster heterogeneity assumptions are incorrect or change over time | • Since key parameters may not remain stable over time (e.g. incidence, coefficient of variation), the study protocol should report the likely power of a cluster-level analysis |

Four entomological outcomes were included in the trials as secondary outcomes, requiring collection by human landing catches and UV light trap. The master protocol determined that entomology outcomes would be collected monthly at a sub-set of clusters. The meta-analysis found a larger ATSB effect size for clinical incidence in the subset of clusters with entomological surveillance than among the full trial population, most likely as a consequence of an imbalance in baseline incidence between arms among the entomological surveillance clusters. While the restricted randomization at each site required an equal number of entomology surveillance clusters in each arm, the procedure did not explicitly require balance among the entomology surveillance clusters. Such requirements may indeed be too restrictive, resulting in a limited number of valid randomization sequences. A trial of spatial repellents in Indonesia reported a four-fold larger effect of the intervention in the subset of entomology surveillance clusters (selected based on the highest human landing rate), and the authors report achieving balance in the entomology surveillance clusters [31]. Where resources are insufficient for comprehensive entomology monitoring in all trial clusters, reducing the frequency or intensity of sampling per cluster is preferable to restricting entomological data collection to a subset of clusters. This strategy was applied in recent insecticide-treated net trials in which outcomes were collected in all clusters, but entomological sampling was generally limited to one trap method, reduced trapping frequency, or reduced the number of traps used in each cluster [32–34].

The ATSB trials’ primary entomological indicator was parity. Since ATSBs represent a new vector control paradigm, an expansive approach was taken in selecting entomological indicators to monitor in the trials. However, parity has limitations as a trial outcome. Firstly, the proportion of female mosquitoes found parous can be high and dynamic in high transmission settings [35], limiting feasible effect sizes and the sensitivity of this outcome. Secondly, parity does not directly correlate with transmission, since mosquitoes must have reached at least their second gonotrophic cycle to transmit *Plasmodium*. Limited availability of baseline entomological data due to COVID-19 restrictions was a further challenge in the design and analysis of the Phase III trials.

The finding of an interaction in Kenya between the effect of Westham Sarabi ATSB on incidence and the predominant vector species in the cluster was unexpected, and subgroup analyses by vector species are challenging to interpret. Baseline data to confirm vector species present at each site may have permitted the very high *An. funestus* abundance clusters to be balanced between arms. Indeed, entomological outcomes were noisy, and our observations of slight and not statistically significant increases in parity and sporozoite positivity and decreases in abundance and landing rate in the intervention arm compared to control are thought to be a chance finding, since age-dependent feeding on ATSB by mosquitoes would be very unlikely to result in this pattern of findings.

The trials in Kenya, Mali, and Zambia were designed according to contemporary best practice, with master protocol and statistical analysis plan reviewed prior to the start of the trials and subsequently published [14, 18] but there were nevertheless limitations. First, in an effort to move rapidly towards the opening of ATSB as a new product class, it was decided to conduct the three trials concurrently, rather than sequentially. Particularly for a new intervention with limited preliminary data or assessment of deployment strategies, operating concurrent trials limited the opportunity to build field experience across sites or to use interim results to inform protocol modifications. A sequential approach was valuable in the early ITN trials, where the sample size for the western Kenya trial was expanded following results from Burkina Faso, which indicated that a 30% effect size may not be attainable [36]. Additional small-scale Phase II studies across multiple sites may also have provided further information on the Westham Sarabi ATSB mechanism of action, reducing the risk of moving immediately to large-scale trials to confirm efficacy, particularly for interventions that are the first in a new product class. While the Phase III trials were costly and found that the deployment approach for the Westham Sarabi product did not result in expected reductions in epidemiological or entomological outcomes, the cost of evidence generation through such trials should be balanced against the cost of scaling interventions where efficacy is not known but preliminary studies are promising and mathematical models indicate the intervention may have substantial benefits. Research partners and funders must balance a complex set of priorities and risks, weighing the desire to move promising products to market quickly so they can avert cases sooner, against taking a slower approach to systematically build the evidence base.

Cluster randomization is recommended in trials where interventions by their nature are delivered to groups rather than individuals and where the intervention offers both direct and indirect benefits. The coefficient of variation, a measure of variability between clusters, is a critical parameter in determining the number of clusters required for adequate power to detect the desired difference in outcome between arms. Ideally, coefficient of variation is calculated using baseline data from the study area, however our results indicate that CV may not remain stable between the baseline and trial periods, adding further complexity to the use of this parameter in sample size estimation. Using model-based approaches to estimate the coefficient of variation and a 95% confidence interval is a potential solution [28], providing information on the likely range of this parameter. Baseline incidence estimates from the Kenya and Zambia trial sites highlighted an additional challenge: estimating CV from baseline data when some clusters report limited person-time at risk due to operational constraints, resulting in imprecise baseline cluster incidence estimates and zero-incidence clusters. The Sarabi ATSB trial results also illustrate the inherent difficulties in assessing efficacy at sites with high heterogeneity. The western Kenya site had a substantially higher coefficient of variation than anticipated at protocol development and it was higher during the trial than at baseline. As the coefficient of variation increases above 0.5, cluster randomized trials become rapidly infeasible due to the very large number of clusters required. For example, the Kenya site would have required more than 100 additional clusters (176 total) to detect a 30% effect size with 80% power under the observed coefficient of variation of 0.75. As the number of clusters increases, the study site also increases in size, potentially further increasing the level of heterogeneity between clusters. When a high coefficient of variation is found at baseline, researchers may need to either reconsider their study site and design, or consider if adequate minimum information would be available in a “worst case” scenario where analysis is limited to cluster-level summaries due to high intra-cluster correlation and high between cluster variance.

While there are instances in which vector control trials have successfully demonstrated public health benefits through changes in clinical incidence in high transmission settings [33, 37, 38], current malaria intervention efficacy trials are designed to measure the benefit of the intervention in a setting where both arms have access to the standard of care interventions, most commonly ITNs. This design presents an additional hurdle in evaluation of new interventions, since they must demonstrate an additional benefit on top of that provided by existing standard of care tools. The effect sizes of malaria interventions tested in cRCTs have decreased since the 1990s, likely a consequence of the layering of multiple interventions [39]. Non-inferiority trials offer an alternative approach where it is possible to directly compare two different interventions rather than the incremental effect of a specified intervention. However non-inferiority trials are likely unethical for new or first-in-class interventions that do not have prior efficacy data, due to the need to maintain equipoise.

## Conclusions

This meta-analysis did not provide any conclusive evidence that the Westham Sarabi v1.2 ATSB deployed at a rate of two devices per residential structure results in statistically significant changes in any of the monitored epidemiological or entomological outcomes. However, there was evidence for a dose-response relationship between increasing spatial density of Westham Sarabi ATSB and reductions in clinical malaria incidence, after accounting for the observed coverage of Sarabi devices that remained in good condition (increasing the number of ATSBs per hectare by 10 was associated with 19% reduction in incidence). These results suggest that using the current approach of ATSB devices placed on structure walls, the most suitable environments for deployment may be urban or peri-urban settings where such high spatial densities can be achieved. Further research exploring alternative deployment approaches for ATSB products may be merited to determine the spatial density required for an observable effect, along with refinement of ATSB product design to enhance attractiveness, mosquito feeding rates, and physical durability.

## Acknowledgements

We wish to thank the many individuals whose involvement in the Phase III trials in Kenya, Mali, and Zambia enabled this meta-analysis to be conducted. This includes the participating communities, data collection teams, laboratory staff, local, regional, and national health authorities, and ATSB trial implementation and management teams at each site. The findings and conclusions contained within are those of the authors and do not necessarily reflect positions or policies of the US Centers for Disease Control and Prevention, IVCC, the Bill & Melinda Gates Foundation, SDC or UK Aid.

## Data availability statement

Data used to generate results reported in this manuscript are available freely available through Figshare, https://doi.org/10.6084/m9.figshare.32030076.

